# MIMIC in the OMOP Common Data Model

**DOI:** 10.1101/2020.08.14.20175141

**Authors:** Nicolas Paris, Adrien Parrot

## Abstract

**Objectives:** In the era of big data, the intensive care unit (ICU) is very likely to benefit from real-time computer analysis and modeling based on close patient monitoring and Electronic Health Record data. MIMIC is the first open access database in the ICU domain. Many studies have shown that common data models (CDMs) improve database searching by allowing code, tools and experience to be shared. OMOP-CDM is spreading all over the world. The objective was to evaluate the difficulty to transform MIMIC into an OMOP (MIMIC-OMOP) database and the benefits of this transformation for analysts.

**Material & Method:** A documented, tested, versioned, exemplified and open repository has been set up to support the transformation and improvement of the MIMIC community’s source code. The resulting data set was evaluated over a 48-hour datathon.

**Result:** With an investment of 2 people for 500 hours, 64% of the data items of the 26 MIMIC tables have been standardized into the OMOP CDM and 78% of the source concepts mapped to reference terminologies. The model proved its ability to support community contributions and was well received during the datathon with 160 participants and 15,000 requests executed with a maximum duration of one minute.

**Conclusion:** The resulting MIMIC-OMOP dataset is the first MIMIC-OMOP dataset available free of charge with real disidentified data ready for replicable intensive care research. This approach can be generalized to any medical field.

## INTRODUCTION

Intensive care units (ICUs) are designed to provide comprehensive support to the most severely-ill patients within a hospital [1]. Mortality is typically high among these patients, both during and after the hospital stay [2]. Understanding the effectiveness of interventions on patient outcomes remains a challenge, due to heterogeneity of patients, complexity of disease, and variation in care patterns. Intensivists use a limited level of evidence to guide decision making [3] whereas ICUs are a high density environment for data production.

With the increasing adoption of electronic health record (EHR) systems around the world leading to large amounts of clinical data [4] and the development of data mining, innovation throught health data is likely to play an important role in clinical medicine [5]. Indeed, based on important medical informations, expectations are to improve clinical outcomes and practices, enable personalized medicine and guide early warning systems, and also easily enroll a large, multicenter cohort while minimizing costs.

*MIMIC-III* (Medical Information Mart for Intensive Care) is a high granularity dataset of over 60,000 intensive care stays and 46 000 unique patients from two successive ICU systems at the Beth Israel Deaconess Medical Center in Boston admitted from 2001 to 2012. It is the first ICU database available for free and has been intensively used in research resulting in more than 300 international publications. However, its monocentric nature makes it difficult to generalize findings to other ICUs. The MIMIC relational data model reflects the original intensive care information systems, as evidenced by the two separate inputevent_mv and ouputevent_cv [6] or the two separate terminologies for physiological data. This leads analysts (datascientists, statisticians, etc.) to reconcile the corresponding data to address this heterogeneity during the pre-processing step of each study.

For Kahn et al. [7], “databases modelling is the process of determining how data are to be stored in a database”. It specifies data types, constraints, relationship and metadata definitions and provides a standardized way to represent resources/data and their relationships. Some studies have shown that using a common data model (CDM) by standardizing the structure (data model) and concepts (terminological model) of the database allows large scale multicenter research, exploitation of rare diseases or rare events and catalyzes research by sharing practices, source code and tools [8, 9]. However, some studies have shown that the results are not fully reproducible from one CDM to another [10] or from one centre to another [11]. Some approaches argue that keeping the local conceptual model [12] and the local structural model [13] leads to better results. On one hand, keeping MIMIC on its specific form will not solve the limitation for multicenter research but on the other hand, a fully standardized form would introduce other disadvantages, such as loss of data and lower computational performances. The ideal solution is probably in between to allow local or standardized analysis depending on the research question.

*OMOP* (Observational Medical Outcomes Partnership Common Data Model) is a CDM originally designed for multicenter research related to adverse drug adverse events and now extends to medical, labortory and genomic cases. OMOP provides structural and conceptual models relying on reference terminologies such as SNOMED for diagnostics, RxNORM for drugs and LOINC for laboratory results. Several examples of database transformed into OMOP have been published [14, 15] and OMOP stores 682 million patient records from around the world [16]. Each clinical area is stored in different dedicated tables. The OMOP conceptual model is based on a closure table pattern^1^ capable of ingesting any simple, hierarchical and also graph terminologies such as SNOMED. In addition to local terminologies, OMOP defines and maintains a set of standard terminologies to be mapped unidirectionally (local to standard) by implementers. Although OMOP has proven its reliability [17], the concept mapping process is known to have an impact on results [18] and the application of the same protocol on different data sources leads to different results [11]. This shows the importance of keeping local terminologies so that local analysis is still possible. Previous preliminary work has been done on the translation of MIMIC into OMOP [19]. This work remains to be refined and updated for proper evaluation.

In a recent comparative study of different CDM [8, 20] OMOP obtained best results for completeness, integrity, flexibility, simplicity of integration, implementability, for a wider coverage of the structural and conceptual model, a more systematic analysis thanks to an analytical library and to visualization tools and easier access to data through SQL queries. In terms of conceptual approach, OMOP offers a broader set of standard concepts. In terms of structural CDM it is very rigorous in how data should be loaded into specific tables while others CDM such i2b2 are very flexible with a general table that solves all data domains. This rigorous approach is necessary for standardization. Previous work has been done to load MIMIC-III into i2b2 [21] - however the work couldn’t be finalized due to tricky concept mapping to standard terminologies tasks. OMOP has the advantage of not making the terminology mapping step mandatory by keeping the local codes accessible to analysts. Compared to the Fast Healthcare Interoperability Resources (FHIR)^2^, OMOP performs better as a conceptual CDM because the FHIR ressources currently do not specify the terminology to be used for most of the attributes. OMOP relational model can be materialized in csv format and stored in any relational database when FHIR uses json files and needs some processing and higher skills to exploit. Among the above models, OMOP is the best candidate to overcome the MIMIC limitations mentioned earlier.

Our article was guided by the two following dimensions:

1. Data Transformations: evaluate the process of transforming MIMIC into OMOP in terms of time needed, skills required and quality of the result.
2. Data Analytics: evaluate the resulting dataset to support efficient, shareable and real-time analysis.

## 1 MATERIAL & METHOD

### 1.1 Data Transformations

The majority of source code is implemented in PostgreSQL 9.6.9 (Postgres) because it is the primary support for the MIMIC database and allows the community to reproduce our work on limited resources without licensing costs and benefit from recent Postgres improvements in the data processing area. Some elaborated data transformations have been implemented as Postgres functions.

The OMOP CDM version 5.3.3.1 (OMOP) tables were created from the provided scripts with some changes documented in our scripts. OMOP defines 15 standardized clinical data tables, 3 health system data tables, 2 health economics data tables, 5 tables for derived elements and 12 tables for standardized vocabulary. The vocabulary tables were loaded from concepts downloaded from Athena^3^ and the clinical and derived tables were loaded from MIMIC.

MIMIC-III version 1.4.21 (MIMIC) was also loaded into Postgres with the provided scripts. A subset of 100 patients over the 46 000 total MIMIC patients selected based on their broad representativeness in the database and cloned into a second instance to serve as a light and representative development set.

#### 1.1.1 Structural Mapping

The structural mapping aims at moving the MIMIC data into the right place in OMOP with some data transformations. It parts into three phases: conception, implementation and evaluation.

The **conception** phase consists of looping over each MIMIC table and choosing an equivalent location in OMOP for each column. In general both projects were appropriately documented but in several cases, we needed some clarification from MIMIC contributors on the dedicated MIMIC git repository^4^, or from the OMOP community forum^5^. Some trickier choices have been discussed in the MIMIC-OMOP git repository^6^ and can be tracked in the commit logs.

The **implementation** is done by an Extract-Transform-Load (ETL) process which is composed of Postgres scripts, extracting information from the source or concept mapping tables, then transforming and loading an OMOP target table. The scripts are managed sequentially through a main program. In last resort some modification of the structural model of OMOP have been made. A dedicated script recaps all of them. It contains columns name modifications, new columns, columns type modifications or database indexing modification. In particular, each source table has been added a global unique sequence incremented from 0 that serves as the primary key and links in the OMOP target tables. As a result every record is uniquely identified allowing to chain the information with OMOP while simplifying the primary/foreign key maintenance.

Although **evaluating** a structural model is difficult [22], several articles have attempted to assess the quality of the CDM [7, 20]. The criteria developed by Khan et al [23], which refer to the Moody and Shanks metrics [22], have been adapted to assess the quality of the data transformation (table 1).

**Table 1.** Transformation Quality Evaluation Metrics.

| Data Model Dimension | Descriptions |
| --- | --- |
| Completeness - structural mapping | Domain coverage : coverage of sources domains that are accommodated by the standard OMOP model |
| Completeness - conceptual mapping | Data coverage : coverage of sources data concepts that mapped to standard OMOP concept |
| Integrity | "Meaningful data relationships and constraints that uphold the intent of the data's original purpose" [23] |
| Flexibility | The ease to expand the standard model for new datatypes, concepts |
| Integration | The capacity of the standard model to use multiples terminology and links its to standard one |
| Implementability | The stability of the models, the community, the cost of adoption |
| Understandability | The ease of the standard model to be understood |
| Simplicity | The ease of querying the standard model - the model should contains the minimum of concepts and relationship |

Beside Moody and Shanks, we provide a set of controls to guaranty a correct transformation. In order to compare overall statistics, some SQL queries have been setup to compare MIMIC and MIMIC-OMOP and we provide basic populations characterizations. All tables have been covered and tested through simple counts, aggregate counts or distribution checks. We estimated the loss of information during the ETL process by measuring the percentage of both columns and rows lost in the process as other previous studies have done [15]. This is important to note we have chosen not to keep irrelevant informations: for example some rows are known to be invalid in MIMIC or some informations are redundant. Each ETL script has been tested using pgTAP, a unit test framework for Postgres. Each unit test script checked whether a particular OMOP target table was loaded correctly. Integrity constraints (primary keys, foreign keys, non-nullable columns) have been included to apply integrity checks at ETL runtime. The last axe of the structural evaluation is Achilles Sofware. It is an open-source analysis software produced by OHDSI^7^. Like many previous authors, we used the Achilles to assess data quality [24]. This tool is used for data characterization, data quality assessment (Achilles’ heel) and health observation data visualization. It has been common practice to perform Achilles tests and use it as a quality assessment in related studies. All the resulting tables are presented in the results section.

#### 1.1.2 Conceptual Mapping

The conceptual mapping aims at aligning the MIMIC local terminologies to the OMOP’s standard ones. It consists of three phases: integration, alignment and evaluation.

The **integration** phase is about loading both kind of terminologies into the OMOP vocabulary tables. The OMOP terminologies are provided by the Athena tool and were loaded with the associated programs. We have used an export with all terminologies without licensing limitations. The local terminologies have been extracted from the multiple MIMIC tables and loaded in the OMOP concept table. When possible, relevant informations from the original MIMIC tables have been concatenated in the *concept_name* column. MIMIC local concepts were loaded with a *concept_id* identifier starting from 2 billion (lower numbers are reserved for OMOP terminologies^8^). In the OMOP concept table, MIMIC concepts can be distinguished with the *vocabulary_id* identifier equal to “MIMIC code” and a *domain_id* identifier targeting the OMOP table in which the corresponding data is stored. This domain information is used in the ETL to send the information in the proper table. We want to call this method “concept-driven dispatching”. OMOP documentation explains that conceptual mapping has to be done before the structural mapping because the nature of the OMOP standard concepts guides in which table (domain) the information should be stored. The concept-driven dispatching methodology enable changing the concept mapping after the transformation without modifying the underlying ETL code because the latter is dynamically based on the concept table content.

The **alignment** phase to standardizing local MIMIC codes into OMOP standard codes, had four distinct cases. In the *first* case, some MIMIC data is by chance already coded according to OMOP standard terminologies (e.g. LOINC laboratory results) and, therefore, the standard and local concepts are the same. In the *second* case, MIMIC data is not coded in the standard OMOP terminologies, but the mapping is already provided by OMOP (ex: ICD9/SNOMEDCT), so the domain tables have been loaded accordingly. In the *third* case, terminology mapping is not provided, but it is small enough to be done manually in a few hours (such as demographic status, signs and symptoms). In the *fourth* case, terminology mapping is not provided and consists of a large set of local terms (admission diagnosis, drugs). Then, only a subset of the most represented codes was manually mapped. When a manual terminology mapping concept is required, a mapping csv file has been built. This solution can be adapted to medical users who do not have training in database engineering. The spreadsheet has several columns such as local/standard labels, ids and also comments, evaluation metrics and a script loads them into the Postgres when completed. We have chosen to use simple SQL queries that are flexible enough to be queried on demand or to generate a pre-filled csv with the best matches. It uses Postgres full-text ranking features and links local and standard candidates with a rating function based on their labels. This work was performed under the control of an intensivist.

The **evaluation** phase was both quantitative and qualitative. The quantitative evaluation measures the completeness of our work: the percent of concepts that are mapped to a standard. The qualitative evaluation evaluates the correctness. For newly generated mappings it has consisted of manually tagging each mapping with a score between 0 and 1 and eventually write a commentary on each mapped concept. In case the mapping of was provided by OMOP - automatic OMOP terminologies mapping -, the evaluation was performed on a subset of concepts manually picked within each terminology.

### 1.2 Data Analytics

Beyond the model transformation and respect of the OMOP standardisation process we applied some analysis.

MIMIC provides a large number of SQL scripts for preprocess and normalize data, calculate derived scores and defined cohorts as known as “contrib”. Some of them have been implemented on top of the OMOP format to load the OMOP derived tables.

A set of *general denormalized* tables has been built on top of the original OMOP format that have the *concept_name* related to the *concept_id* columns. The concept table is a central element of OMOP and, therefore, it is involved in many joins to obtain the concept label. By precalculating the joins with the concept tables, the denormalized tables faster calculation and simplify SQL queries.

In addition, a set of *specialized analytical tables* has been built on the original OMOP format. The OMOP microbiologicalevents table is a reorganization of the measurement table data of microorganisms and associated susceptibility testing antibiotics and is based on the MIMIC microbiologicalevents table. The OMOP icustays table allows to quickly obtain the patients admitted in resuscitation and is inspired by the MIMIC icustays tables.

The OMOP note_nlp table was originally designed to store final or intermediate derived informations and metadata from clinical notes. When definitive, the extracted information is intended to be moved to the dedicated domain or table and then reused as regular structured data. When the information is still intermediate, it is stored in the note_nlp table and can be used for later analysis. To populate this table, we provided two information extraction pipelines. The *first* pipeline extracted numerical values such as weight, height, body mass index and left ventricular cardiac ejection fraction from medical notes with a SQL script. The resulting structured numerical values were loaded into the measurement or observation tables according to their domain. The *second* pipeline *section extractor* based on the apache UIMA framework divides notes into sections to help analysts choose or avoid certain sections of their analysis. Section templates (such as “Illness History”) have been automatically extracted from text with regular expressions, then filtered to keep only the most frequent (frequency > to 1%).

A 48-hour open access datathon^9^ was set up in Paris AP-HP (Assistance Publique des Hopitaux de Paris) in collaboration with the MIT once the MIMICOMOP transformation was ready for research. This datathon was organised to evaluate OMOP as an alternative data model for accessing and analysing MIMIC data during a real event. Scientific questions had been prepared in an online forum where participants could introduce themselves and propose a topic or choose an existing one. OMOP has been loaded into apache HIVE 1.2.1 in ORC format. Users had access to the ORC dataset from a web interface jupyter notebooks with python, R or scala. A SQL web client allowed teams to write SQL from presto to the same dataset. The hadoop cluster was based on 5 computers with 16 cores and 220GB of RAM memory. The MIMICOMOP dataset has been loaded from a Postgres instance to HIVE thought apache SQOOP 1.4.6 directly in ORC format. Participants also had access to the Schemaspy database physical model to access the OMOP physical data model with both table/column comments and key primary/foreign relationships materializing the relationships between the tables. All queries have been logged.

## 2 RESULT

All transformation processes are freely accessible to the public via the MIMIC-OMOP git repository maintained by MIT-LCP [6]. The repository is based on git and is designed for sharing, improvement, collaboration and reproducible work. Moreover, it is archived on a universal and durable software archive solution^10^. The git repository centralizes the various resources of this work such as documentation, source code, unit tests, as well as questioning examples, discussions and problem issues. It also indicates web resources such as the physical data model for MIMIC^11^ and OMOP^12^ datasets and the Achilles’ web client^13^. All the code to create these statistics is provided on the article’s framagit repository (see Repository Work section).

### 2.1 Data Transformations

The MIMIC to OMOP conversion was performed by two developers (a data engineer and an intensivist) for 500 hours. This includes ETL, git documentation, concept mapping, contributions and unit tests. ETL (with unit tests and generation of ready-to-load archive) on a subset of 100 patients lasts five minutes and enables fast development cycles. The ETL lasts 3 hours to process the whole MIMIC database. The resulting csv archive is almost the same size as the original archive, and MIMIC-OMOP is also the same size as MIMIC once loaded and indexed into Postgres.

#### 2.1.1 Structural Mapping

The result of the Structural Mapping are presented in the table 2. Among of the 37 OMOP tables, the one related to hospital costs were not applicable, some related to derived data were not populated and some tables related to vocabulary were pre-loaded with terminology informations. The 26 tables of MIMIC have been dispatched into 19 OMOP tables. The reduced number of tables results from the differences in design of both models. OMOP stores all the terminologies into one table whereas MIMIC has one table for each terminology and the same applies for facts data that are grouped by nature in OMOP while MIMIC tables are more specialized and respects the source EHR’s design. For example the measurement gather measured information and combines 4 source tables resulting in 365 181 104 rows which is 20% more than the largest MIMIC table. To some extends this is a regression in terms of performances. Two important tables are provided by OMOP to represent the relationship between the data: concept_relationship and fact_relationship. We used them to bind the drugs into a solution, for microbiology / antibiograms and for visit_detail / caresite links. The following SQL query (listing 1) shows how a microorganism is linked to its susceptibility test by a fact_relationship and illustrates the *flexibility* of the model. However this flexibility affects the *simplicity* and the performances of the model by increasing the number of joins within SQL queries.

**Listing 1.**
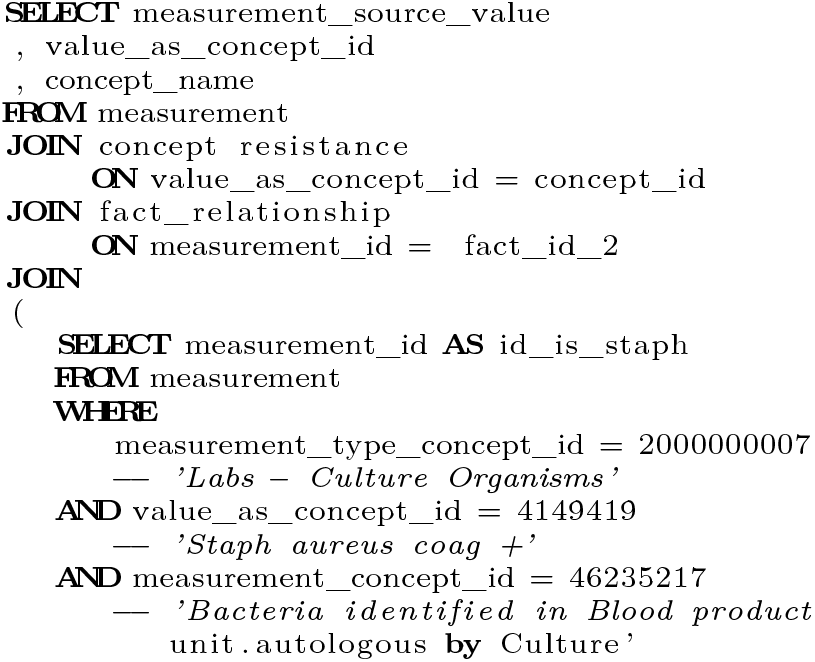
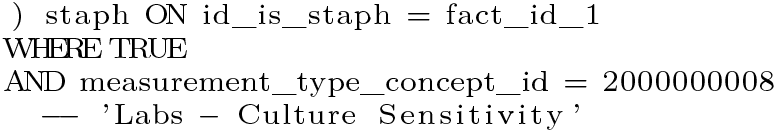
Original table microbiology SQL query

**Table 2.** MIMIC to OMOP data flows.

| OMOP tables | Number of rows | MIMIC tables |
| --- | --- | --- |
| CARE_SITE | 93 | transfers, service |
| COHORT_ATTRIBUTE | 228379 | callout |
| CONCEPT | 30344 | d_cpt, d_icd_procedures, d_items, d_labitems |
| CONDITION_OCCURRENCE | 716595 | admissions, diagnosis_icd |
| DEATH | 14849 | patients, admissions |
| DRUG_EXPOSURE | 24934751 | prescriptions, inputevents_cv, inputevents_mv |
| MEASUREMENT | 365181104 | chart/lab/microbiology/in/output events |
| NOTE | 2082294 | noteevents |
| NOTE_NLP | 16350855 | noteevents |
| OBSERVATION | 6721040 | admissions, chartevents, datetimeevents, drgcodes |
| OBSERVATION_PERIOD | 58976 | patients, admissions |
| PERSONS | 46520 | patients, admissions |
| PROCEDURE_OCCURRENCE | 1063525 | cptevents, procedureevents_mv, procedure_icd |
| PROVIDER | 7567 | caregivers |
| SPECIMEN | 39874171 | chartevents, labevents, microbiologyevents |
| VISIT_OCCURRENCE | 58976 | admissions |
| VISIT_DETAIL | 271808 | admissions, transfers, service |

The table 3 presents the basic characterization of the MIMIC-OMOP population and assesses the overall quality of structural mapping. Fortunately most statistics remain similar between the two versions with still few differences. The table 3 shows MIMIC contains 61,532 intensive care stays while OMOP contains 71 576 intensive care stays. This represents a 16% increase in stays. By desigh MIMIC aggregates information from various systems. Thus the transfer information is divided into several tables, such as admissions, transfers and also icustays. Rather OMOP centralizes this information in the detail of the visit_detail. We also added emergency stays as a normal location for patients throughout their hospital stay (unlike what had been done by MIMIC). The Icustays MIMIC table has not been transformed because it derives from the transfer table^14^ and we decided to assign a new visit_detail row for each ICU stay (based on the transfer table) while MIMIC preferred to assign a new ICU stay if a new admission occurs > 24h after the end of the previous stay.

**Table 3.** Baseline characteristics MIMIC versus OMOP items MIMIC.

| items | MIMIC | MIMIC-OMOP |
| --- | --- | --- |
| Persons (Number) | 46.520 | 46.520 |
| Admissions (Number) | 58.976 | 58.976 |
| Icustays (Number) | 71.575 | 61.532 |
| Gender, Female (Number, %) | 20.399 | 20.399 (43 %) |
| Age (Mean) | 64 years, 4 months | 64 ans, 4 months |
| 0-5 | 8110 | 8110 |
| 6-15 | 1 | 1 |
| 16-25 | 1434 | 1434 |
| 26-45 | 5962 | 5962 |
| 46-65 | 17375 | 17375 |
| 66-80 | 15793 | 15793 |
| >80 | 10301 | 10301 |
| Emergency | 42071 | 42071 |
| Elective | 7706 | 7706 |
| Surgical patients | 19246 | 19246 |
| Length of stay, hospital (median) | 6.46 (Q1-Q3 : 3.74 - 11.79) | 6.59 (Q1-Q3 : 3.84 - 11.88) |
| Length of stay, ICU (median) | 2.09 (Q1-Q3 : 1.10 - 4.48) | 1.87 (Q1-Q3 : 0.95 - 3.87) |
| Mortality, ICU (Number, %) | 5814 (9%) | 5815 (9%) |
| Mortality, hospital (Number, %) | 4511 (7%) | 4559 (6%) |
| Lab measurements per admissions (mean) | 478 | 678 |
| Procedures per admissions (mean) | 4.6 | 4.6 |
| Drugs per admissions (mean) | 82.8 | 82.8 |
| Exit dignosis per admissions (mean) | 11.0 | 11.0 |

This table also shows an increase of the number of laboratory measurements per admission. This is because MIMIC-OMOP gathers laboratory data from both the MIMIC dedicated laboratory table and the chartevents table which is usually not considered for this purpose. For laboratory tests we put a specimen (i.e. a blood sample) for many laboratory results (because one blood sample can be used for several tests), we decided to create as many rows of samples as laboratory tests because the information is not present in MIMIC. The same was true when date information was not provided (*start /end_datetime* for drug_exposure).

As mentioned in the table 4, from 20% to 80% of the source columns has not been kept. Almost all were redundant with others or provided derived information. The main concern is the loss of some timestamps. For example, the MIMIC chartevents tables provides the *storetime* and *charttime* columns, but OMOP only provides one column to store timestamp. Thus, MIMIC *storetime* column was eliminated during the ETL which has been considered the less valuable.

**Table 4.** Data lost.

| Relations | Rows lost | Columns lost |
| --- | --- | --- |
| admissions | - | 30% |
| callout | - | 80% |
| caregivers | - | 50% |
| chartevents | 0.04% | 40% |
| cptevents | - | 60% |
| datetimeevents | 0.0001% | 50% |
| diagnoses_icd | - | 20% |
| drugcodes | - | 60% |
| inputevents_cv | - | 41% |
| inputevents_mv | 10,00% | 46% |
| labevents | - | 34% |
| microbiologyevents | - | 30% |
| noteevents | 0.04% | 19% |
| outputevents | - | 39% |
| patients | - | 50% |
| prescriptions | - | 16% |
| procedureevents_mv | 3,00% | 70% |
| procedures_icd | - | 40% |
| services | - | 34% |
| transfers | - | 47% |

As mentioned in the methods the incorrect entries are not kept in the process. According to the tables 4, four MIMIC tables (inputevents_mv, chartevents, procedureevents_mv, note) have deleted rows in the ETL process. All of them were tagged in MIMIC as erroneous or cancelled.

A set of minor modifications of the OMOP tables structure was made in order to fit the data. All character columns with limited length have been modified to unlimited length since it could cause unpredictable truncation of content, while having no negative impact on Postgres storage size or performance. The visit_occurrence and the visit_detail tables have been corrected accordingly to some discussions on the OHDSI forum. The nlp_note table has been extended with fields mentioned in online documentation but forgotten in the scripts. In addition the *offset* column has been divided into two integer type columns because the offset term is a SQL reserved word and it makes sense to fill the resulting *offset_begin* and *offset_end* resulting columns with integer values.

All the PgTAP unit tests passed. Moreover OMOP had a 100% match of the integrity constraints and the foreign key relationships of the data models. After 18 hours of computations Achilles Heel issued 15 errors, 18 warnings and 8 notifications. This result is good compared to other studies [24].

#### 2.1.2 Conceptual Mapping

The results of the Conceptual Mapping’s completeness are presented in the table 5.

**Table 5.** Terminology Mapping coverage.

| Omop tables (domain) | Records | % Mapped records | Concepts source | % Mapped concepts source |
| --- | --- | --- | --- | --- |
| CARE_SITE | 144 | 100% | 58 | 100% |
| CONDITION_OCCURRENCE | 716595 | 90% | 6984 | 94% |
| DRUG_EXPOSURE | 24934751 | 38% | 7398 | 56% |
| MEASUREMENT | 40141521 | 73% | 1035 | 76% |
| OBSERVATION | 6721040 | 68% | 1440 | 80% |
| PERSONS | 93040 | 100% | 43 | 100% |
| PROCEDURE_OCCURRENCE | 1063525 | 99% | 2203 | 99% |
| SPECIMEN | 39874171 | 70% | 92 | 77% |
| NOTE | 2082294 | 100% | 15 | 100% |
| VISIT_OCCURRENCE | 176928 | 100% | 34 | 100% |
| VISIT_DETAIL | 396932 | 100% | 28 | 100% |

We have often mapped many source concepts to a unique standard *concept_id* because MIMIC provides a large number of equivalent concepts. For example MIMIC provides 6 distinct concepts for body temperature: Temperature C, Temperature C (calc), Temperature F, Temperature F (calc), Temperature Fahrenheit and Temperature Celsius. All of them have been mapped to the LOINC “Body temperature” and numerical values have been normalized.

OMOP’s terminology coverage has already been rated as excellent [20]. We used the OMOP terminology mappings - NDC-RxNorm, ICD9-SNOMED, CPT4-SNOMED - to standardize a consequent set of MIMIC non-standard terminologies.

The automatic OMOP terminologies mapping was evaluated by an intensivist. This results are in favor of a good *integration* of the model. We checked 100 elements for each mapping used (NDC, ICD9 and CPT4). ICD9 and CPT4 are correctly mapped to SNOMED (100%). But only 85% of NDCs are linked to a correct RxNorm code. This is partly due to an incorrect NDC drug code (from MIMIC), partly because only 78% of NDC codes are mapped to Rxnorm. Moreover, even if this does not seem to have affected our ETL we know that some of ICD-9-CM codes can have a one-to-several match with SNOMED^15^ (28%).

In several cases, OMOP had no suitable concepts for the ICU specific cases. In particular, the visit_detail table does not yet introduce relevant information and duplicates information from visit_occurrence table. Therefore we extended the concepts to track bed transfers and room transfers thought *admitting_concept_id, discharge_to_concept_id* or *visit_type_concept_id* columns. These added concepts have been introduced with *concept_id* between 2 billion and 2.001B to distinguish them with OMOP concepts (0 to 2B) and MIMIC locals (>2.001B).

Some local concepts could not mapped to standard ones. This unmapped concepts are linked with the *concept_id* = 0 and appear in different cases. In the first case, the local concept has no equivalent in the standard concept set. In the second case, it has not yet been mapped and may have a standard equivalent. In the third case, the value is missing and cannot be mapped. In our opinion, although not all of these cases can be used for standard queries, they should have a different concept identifier in order to be treated differently (not only *concept_id* = 0). Some of the *domains_id* do not match the table name, it makes sense because the observation domain can be measurement table and vice versa. Although various types of information are stored in the measurement table, the dedicated OMOP concepts for the *measurement_type_concept_id* column were not sufficient to distinguish them. Therfore we added some (Labs - Chemistry, Labs – Culture Organisms etc).

### 2.2 Data Analytics

Some MIMIC raw informations have been transformed and added to meet the structural model. The laboratory textual values have been splitted into operators, numeric values, and units when needed with a dedicated Postgres stored procedure. The free text conditions have been normalized and mapped to OMOP standard codes to meet the conceptual model.

As indicated in the methods section, we have provided many *derived values*. Common derived informations were introduced and loaded: corrected serum calcium, corrected serum potassium, P/F ratio, corrected osmolarity, SAPSII.

*Denormalized derived* tables improve SQL query performances and verbosity. In addition, the resulting tables are much more human readable with the concept label directly in table and greatly reduces joins. Therefore, a little denormalization greatly improves the analysts experience of the data model and the simplicity by adding some redundancy in the data while not interrupting existing SQL queries. Moreover, these normalized views are backward compatible and remain standardized allowing the creation of multicentric algorithms. We provide two examples of *materialized specialized views* derived from microbiologyevents and icustays MIMIC that simplify the experience for scientists (listing 2). This results reflect the lack of *simplicity* of the model in its original form but this can be easily overcome with such analytics tables.

**Listing 2.**
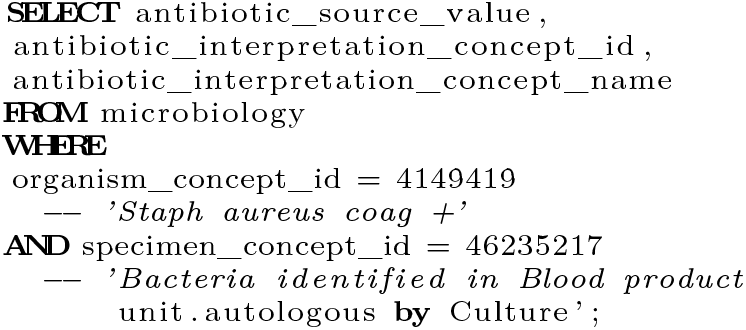
Optimized and denormalized microbiology table SQL query

This results are in favor of a good *flexibility* of the model allowing to store derived data.

The note section extraction pipeline resulted in 1200 sections that were collected and then manually filtered to exclude false positives. 400 similar groups were highlighted. The extracted sections have not been mapped to standard terminologies such as LOINC Clinical Document Ontology (CDO). The reason for this is that the CDO LOINC decided not to maintain their sections from its standard, considering that these sections were not widely used^16^.

The French Hospitals of Paris (AP-HP) organized a datathon with MIMIC-OMOP. 25 teams, 160 participants had 48 hours to undertake a clinical project using the database MIMIC-OMOP through 15,000 requests with a maximum duration of one minute. They had the opportunity to create mixed teams: clinicians brought the issues that required data mining, as well as their data expertise; data scientists judged the technical feasibility and finally implemented the various analyses needed. Writing standard queries (i.e. with standard concepts) requires knowing the organization of relational models (SQL) and also mastering the graphical nature of certain terminologies such as SNOMED-CT in order to capture all potential codes that might be related to the one analysts think of first. Overall the teams quickly mastered the OMOP model and managed to produce results at the end of the datathon. This results are in favor of a good *understandibility* and *simplicity* of the model.

## 3 DISCUSSION

### 3.1 Data Transformations

The choice of a simple SQL based ETL over a dedicated ETL software has several advantages. SQL as the unique language factors both people’s knowledge and computer resources allowing analysts to become implementers and revise code or contribute to transformations. SQL was also used for the semantic mapping and we did not use language algorithm has proven to be effective [25] and OHDSI provides USAGI^17^. The use of csv as format for sharing informations is simple and universal. Both are standard and target a large community (physicians, engineers and analysts) with translational profiles and is compatible with multiple technology.

The calculation time of the ETL on the Postgres instance on a modest personal computer is compatible with a community work where the collaborator can clone the source code and configure a development instance to reproduce or improve the work.

By choosing a public git repository for documentation and source code support, this allows analysts to learn more about the project and learn how to contribute.

The highly active OMOP forum is full of details and in training. It contrasts the implementation guide suffers from not being as detailed and maintained. We believe that the OMOP community would greatly benefit from a systematic and concise synchronization between the forum, mailing lists, source code repository and end user documentation.

Any data transformation is likely to generate bugs that can have a later impact in medical research. The foundations of the Relational database management system (RDBMS), such as transactions, standardization and integrity constraints, are integrated safeguards that have been useful throughout the process. In addition the implemented unit tests ensure that past bugs are behind us. An ideal but complex validation method [26] would be to replicate existing MIMIC studies and ensure that the results are consistent across data models. OHDSI Achilles tool completes our quality assessment. It it is a surprisingly slow tool to process. The rules and their descriptions are difficult to understand. More specific one should be provided and described.

Another missing aspect is a set of quality tables for assessing and measuring data quality. MIMIC have a column to keep track of corrupted information. It would be interesting to be able to keep the disordered data in OMOP and enable research in the data cleaning/quality field. Although OMOP-CDM provides rules to name columns, there are some mistakes and we have to modify it. One the first hand, it is a problem for a CDM to contain errors but this other hand it is easy to relay issues that are now corrected.

### 3.2 Data Analytics

It is important that OMOP maintains a level of standardisation in order to simplify ETL and make it consistent. However, once done, it makes sense to give access to scientific data through more denormalized and specialized tables. There are many concerns about OMOP’s performance and optimization. However, there will never be a perfect multi-purpose case table, and it is the responsibility of the data scientists to build his own, simplified, specialized tables for his research and to respond effectively and clearly to his needs.

The derived data integrate quite well into OMOP. We used note_nlp to store information derived from notes, measurement to store derived numerical informations and cohort_attribute to store derived scores. However, it is not yet clear whether derived data should be stored by domain or whether it should be stored in dedicated derived tables. We found that there are no tables to track the source and description of these data.

The pipeline notes’ section extractor we used was based on apache UIMA framework. While some methods already exist to extract medical sections [27], the prior work of describing sections was too high, and we opted for a naive approach.

Last but not least, as noted in the introduction, a good CDM for the ICU would allow for near real-time early warning systems and inference modelling on fresh data. OMOP is clearly designed to provide a static data set and does not have real-time ingestion and data versioning control mechanisms like EHR usually do. Analysis of static data sets is essential for reproducible results. However, when the algorithms need to be moved to the bed side, it is necessary to have fresh data and a way of re-identifying the patient that OMOP does not yet provide. That said, a solution like HL7 FHIR is a great way to implement real-time inference from EHR data, and that’s how FHIR and OMOP are complementary. This has already been studied^18^ but needs further optimisation.

The datathon showed that distributed platforms with basic hardware provide SQL tools for Online Analytical Processing (OLAP) with excellent performances that overcome RDBMS weaknesses. Therefore, it takes advantage of SQL language analysis functions such as grouping, windowing, assembling and mathematical functions that are often missing in NoSQL databases. While some are open-source, those distributed technology are not easily accessible but cloud based solution are more and more affordable for researchers.

The real life test of the datathon revealed the strong need to make the physical data model accessible, including comments on columns and tables, and we discovered that an open-source tool called schemaspy was very helpful. In addition, we found that the git repository is the best place to document and interact with the community.

## 4 CONCLUSION

The data transformations of MIMIC in OMOP model is a success. The transformation of MIMIC into OMOP has required efforts that remain reasonable. It is and always will be a work in progress because standard concept mapping is an almost infinite process with constant improvements. Fortunately, the published version of MIMIC-OMOP is search-ready and already offers the same scope of data as the original MIMIC version and even more with the derived data. It is publicly available on the git repository and have been designed to be easily revised, copied or enriched according to the OMOP or MIMIC philosophy by any users who knows SQL.

The OMOP model is powerful because it allows a broad spectrum of analysis from specialized local models to evidence-based statistical analysis in an easy-to-learn and accessible format. The major complexity of this model is intrinsically linked to terminologies complexity with the use of its closure table [28].

Compared to the original MIMIC data model, working with OMOP offers the ability to write standard code and analyses that could benefit other international users.

As we have seen, the effectiveness of the OMOP model has some weaknesses because it seems to focus on consistency rather than performance. However, we have shown that it is easy to overcome the weaknesses and improve OMOP with a set of design or technology optimization and a dedicated structure that ultimately remains a standard and shareable because it derives from the original model.

The *first* major contribution of this study is to evaluate OMOP in the context of a freely accessible and well known database - MIMIC. The *second* major contribution is to provide a freely accessible dataset in the OMOP format that could be useful to researchers. The *third* major contribution is to share with the OMOP community some useful transformations dedicated to intensive care that can be reused on any OMOP data set.

Future studies on the evaluation of structural and conceptual mapping through practical research studies on local and standard coding will be carried out. In addition, we plan to enhance the USAGI OHDSI concept mapping tool to enable international concept mapping suggestion to transform other foreign ICU databases. Finally, research on how to articulate FHIR and OMOP to get the best of both worlds (information at the patient level versus information at the multicenter level) and improve bedside care have to be done.

## 5 GRANT

This research received non specific grant from any funding agency in the public, commercial, or not-forprofit sectors

## 6 REPOSITORY WORK

All the latex files, statistics, pdf of the article are provide online: https://framagit.org/aphp/mimicomop-article.

## Data Availability

The mimic data are available on physionet.org. The transformation to omop is available as program.

## Footnotes

1 https://karwin.blogspot.com/2010/03/rendering-trees-with-closure-tables.html

2 https://www.hl7.org/fhir/

3 https://www.ohdsi.org/analytic-tools/athenastandardized-vocabularies/

4 https://github.com/MIT-LCP/mimic-code

5 http://forums.ohdsi.org/

6 https://github.com/MIT-LCP/mimic-omop

7 https://www.ohdsi.org/web/achilles/

8 http://www.ohdsi.org/web/wiki/-doku.php?id=documentation:cdm:concept

9 http://blogs.aphp.fr/dat-icu/

10 https://www.softwareheritage.org/

11 https://mit-lcp.github.io/mimic-omop/schemaspymimic

12 https://mit-lcp.github.io/mimic-omop/schemaspyomop

13 https://mit-lcp.github.io/mimic-omop/AchillesWeb

14 https://mimic.physionet.org/mimictables/icustays/

15 https://www.nlm.nih.gov/research/umls/mapping_projects/icd9cm_to_snomedct.html

16 https://loinc.org/news/loinc-version-2-63-and-relmaversion-6-22-are-now-available/

17 https://github.com/OHDSI/Usagi

18 http://omoponfhir.org/

